# Estimating Lower Bounds for COVID-19 Mortality from Northern Italian Towns

**DOI:** 10.1101/2020.06.10.20125005

**Authors:** Thomas S. Coleman

## Abstract

For COVID-19 the Infection Fatality Rate or IFR – a crucial variable in epidemiological modeling – is difficult to estimate because many cases are asymptomatic and the overall infection rate is generally not known. Circumstances in the Italian provinces of Milano, Bergamo, Brescia, and Lodi allow estimation of lower bounds for age- and sex-specific all-cause excess mortality (a proxy for IFR) since anecdotal reports indicate some towns were close to fully infected. Using data from ISTAT on mortality from January 1 through April 15 for 2020 and the three preceding years, I estimate excess mortality by sex and age categories (0-14, 15-54, 55-64, 65-74, and 75+ years) while controlling for town-specific mortality that proxies for town-specific infection rate. The 99th percentile from the tail of the town distribution gives a lower-bound estimate for COVID-19 mortality. The overall population-weighted mortality at the 99th percentile is 1.09 percent (95% CI 1.06-1.14). The age- and sex-specific rates vary considerably: for men age 65-74 the estimate is 2.10 percent (95% CI 1.94-2.28) which is 3.5-times higher than men 55-64 and 2.7-times higher than women 65-74.

## 1 Summary of Results

Towns in the four northern Italian provinces of Milano, Bergamo, Brescia, and Lodi were heavily infected with the coronavirus in the first few months of 2020. Some towns had substantial excess mortality:

- Nembro, 147 extra deaths, population 11,526, excess mortality 1.275 percent
- Alzano Lombardo, 102 extra deaths, population 13,655, excess mortality 0.75 percent

If the towns had been fully infected during this period the excess mortality would provide an estimate of mortality due to COVID-19. Since the towns may have been less than fully infected these provide lower bounds.

The excess mortality provides a valuable bound for COVID-19 mortality and it would

Generalizing the idea to multiple towns provides better precision in the estimates, and opens the way for estimating mortality by age and sex. A Poisson count model provides the statistical framework to pool data across towns, estimating age- and sex-specific mortality rates, while controlling for differences across towns in the degree of excess mortality. There are 612 towns with deaths by age and sex available from ISTAT for January 1 through April 15 for the years 2017-2020 in the northern provinces of Milano, Bergamo, Brescia, and Lodi – a population of 5,753,296 or 98 percent of the regional population.

Town-specific random effects control for differences across towns in the level of excess mortality which presumably results from differences in infection rates, and also provide a method for estimating mortality for the most-infected towns. The upper tail (99th percentile) of the town distribution corresponds to the highest-mortality towns – presumably those most infected by the coronavirus. The 99th percentile out of 612 towns is roughly the 606th town, so at the upper tail but not simply the most-infected town.

Table 1 shows overall excess mortality of 1.09 percent (95% CI 1.06-1.14),^1^ an IFR higher than many published estimates (e.g. Ioannidis [2020] (surveying published studies) reports 0.02 percent to 0.40 percent; CDC [2020] uses an “upper bound” for disease severity of *case* fatality rate 1 percent and asymptomatic case percentage of 50 percent, implying an IFR of 0.5 percent; Grewelle and Leo [2020] does estimate a higher global IFR (1.04 percent), but this is equal to our lower bound). The IFR shows a dramatic age and sex profile: men age 65-74 have excess mortality 12-times higher than younger men and 2.7-times higher than women of the same age. The overall sex ratio of male-to-female excess mortality (which is close to one) masks the high age-specific sex ratios because the Italian

**Table 1:** Estimated Excess Mortality, Approximate 99th Percentile of Town Infection Distribution

| Age | Male | 95% CI | Female | 95% CI | Total | 95% CI | Ratio M/F | 95% CI |
| --- | --- | --- | --- | --- | --- | --- | --- | --- |
| 0-14yr | 0.01% | 0.01-0.02 | 0.01% | 0-0.01 | 0.01% | 0-0.01 |  | 0, |
| 15-54yr | 0.06% | 0.05-0.07 | 0.04% | 0.03-0.04 | 0.05% | 0.04-0.06 | 1.65 | 1.40-1.91 |
| 55-64yr | 0.59% | 0.54-0.64 | 0.22% | 0.19-0.25 | 0.40% | 0.37-0.43 | 2.65 | 2.34-3.14 |
| 65-74yr | 2.10% | 1.94-2.28 | 0.80% | 0.74-0.87 | 1.41% | 1.30-1.52 | 2.62 | 2.47-2.80 |
| 75+ | 8.95% | 8.36-9.52 | 6.47% | 6.02-6.89 | 7.42% | 6.93-7.88 | 1.38 | 1.34-1.45 |
| TOTAL | 1.14% | 1.07-1.21 | 1.04% | 0.97-1.11 | 1.09% | 1.02-1.16 | 1.09 | 1.06-1.14 |

Excess mortality rates calculated by ten age-sex categories then weighted to totals using age-specific population in the regions of Milano, Bergamo, Brescia, and Lodi. Non-parametric bootstrap confidence intervals as described in the text.

^2^Applying the age- and sex-specific excess mortality to the U.S. age profile produces a bound for the U.S. overall IFR of 0.73 percent – Table 2.

**Table 2:**
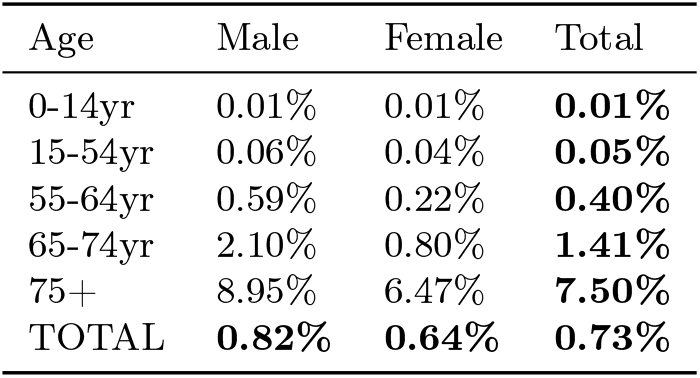
Estimated Excess Mortality with Totals Re-Weighted by US Population, Approximate 99th Per-centile

These estimates of all-cause excess mortality provide particularly firm and useful bounds for COVID-19 mortality because they do not depend on the assumptions often necessary with other measures. They do not depend on ascribing cause of death or determining exposure or proportion asymptomatic infections.

These excess mortality estimates provide lower bounds for COVID-19 IFR in realistic circumstances, but we need to discuss their interpretation. The Infection Fatality Rate (IFR) is a key variable in epidemiological modeling and is usually considered a constant, but in reality IFR can vary with circumstances. There are three channels through which a COVID-19 epidemic will affect mortality. The first is death caused directly by COVID-19 but under “good medical treatment.” We might call this the “clinical” or “best-case” IFR and this is what is usually meant when discussing the IFR. The second is higher mortality still caused directly by COVID-19 but due to ineffective or lesser medical care that may exist during the initial outbreak or disorganized medical conditions. The third is excess mortality from other causes due to disorganized medical treatment, for example cardiac cases not seeking or receiving adequate care – not technically part of the IFR but still mortality resulting from the disease.^3^ The three channels combined we might label the “initial response,” “real-world,” or “worst-case” IFR – due not only to the clinical conditions of infection but also to real-world treatment challenges and the “fog of war” in fighting a new disease.

Reports on the response of hospitals and ICU beds in Lombardy (e.g. Manca [2020], Grasselli et al. [2020]) indicate that the medical system was stressed but not overwhelmed and hospitals responded with large increases in ICU capacity. From anecdotal reports it appears that increased mortality due to the third channel (non-treatment of non-COVID conditions) was small, and due to the second channel (ineffective COVID care) was probably not substantial. Nonetheless, as time, knowledge, and medical practice progresses this initial response IFR will likely decrease.

The estimates in Table 1 provide benchmarks against which to compare parameters used in epidemiological modeling. In this respect it is instructive to examine recent CDC simulations (CDC [2020]). Re-weighting our age-specific estimates for US population (as shown in Table 2) we find 0.73 percent overall and 3.95 percent for age 65+. The CDC scenario 5 (“current best estimates”) assumes 35 percent asymptomatic cases and case fatality rates of 0.4 percent overall, 0.2 percent for 55-64, and 1.3 percent for age 65+, implying IFR of 0.26 percent overall, 0.13 percent for 55-64, and 0.845 percent age 65+. These are substantially lower than the observations from northern Italy and raise the question of whether U.S. mortality will in fact be improved relative to Italy by a factor of roughly three.^4^

By estimating excess mortality using fixed effects for demographic *and* town effects, we can examine town-by-town mortality while still pooling data across demographics and towns. Table 3 shows six towns between the 98th and 99th percentile with the highest predicted overall mortality.^5^ There are a number of towns in the tail of the overall mortality distribution (for exampleGazzaniga, Castiglione d’Adda, and Nembro) that have moderate populations and statistically significant estimated fixed effects. This indicates that the estimates from the tail of the random effects estimator shown in Table 1 are not simply a statistical artifact: Estimates for the upper tail of the random effects estimator and for individual towns tell the same story of high excess mortality for highly-infected towns.^6^

**Table 3:** High Mortality Towns – Actual and Predicted Excess Morality, and Estimated Fixed Effect

|  | Pop | Actual Overall | Predicted Overall | Estimated Fixed Effect value | z-ratio | Actual M55-64 | Actual M65-74 |
| --- | --- | --- | --- | --- | --- | --- | --- |
| Gazzaniga | 5,018 | 1.32% | 1.27% | 1.413 | 8.82 | 0.00% | 2.38% |
| Castiglione d’Adda | 4,646 | 1.29% | 1.27% | 1.554 | 8.74 | 0.28% | 4.68% |
| Valbondione | 1,032 | 1.36% | 1.28% | 1.587 | 4.10 | 3.85% | -1.56% |
| Averara | 181 | 1.10% | 1.29% | 1.583 | 1.73 | 0.00% | 0.00% |
| Nembro | 11,526 | 1.31% | 1.31% | 1.554 | 13.88 | 0.88% | 3.15% |
| Valtorta | 267 | 1.50% | 1.37% | 1.399 | 2.09 | 0.00% | 3.57% |

Towns between roughly the 98th and 99th percentiles in the distribution of overall predicted excess mortality (towns 599 to 604 out of 612 towns)

The “Actual” excess mortality for males aged 55-64 and 65-74 also show why pooling across the towns is valuable. Towns with low population will have substantial random variation, particularly for groups with low mortality. Even if the overall town mortality is enough to provide reliable overall mortality estimates, the age-specific mortality measures may not be reliable. The town of Valbondione provides an example. The town has a population of 606 with only 57 men aged 55-64. With one death in 2019 versus none in 2020 the town has a large negative excess mortality, when in fact the variation in number of deaths is likely due to random chance given the small size of the relevant population.

## 2 Background and Summary Statistics

There are 612 towns (Comunes) having data provided by ISTAT in the four provinces of Milano, Bergamo, Brescia, and Lodi. Table 4 shows summary statistics. The median town population is 4,108. Median observed mortality for 2020 was 0.58 percent, with the median *excess* mortality 0.25 percent.^7^

**Table 4:**
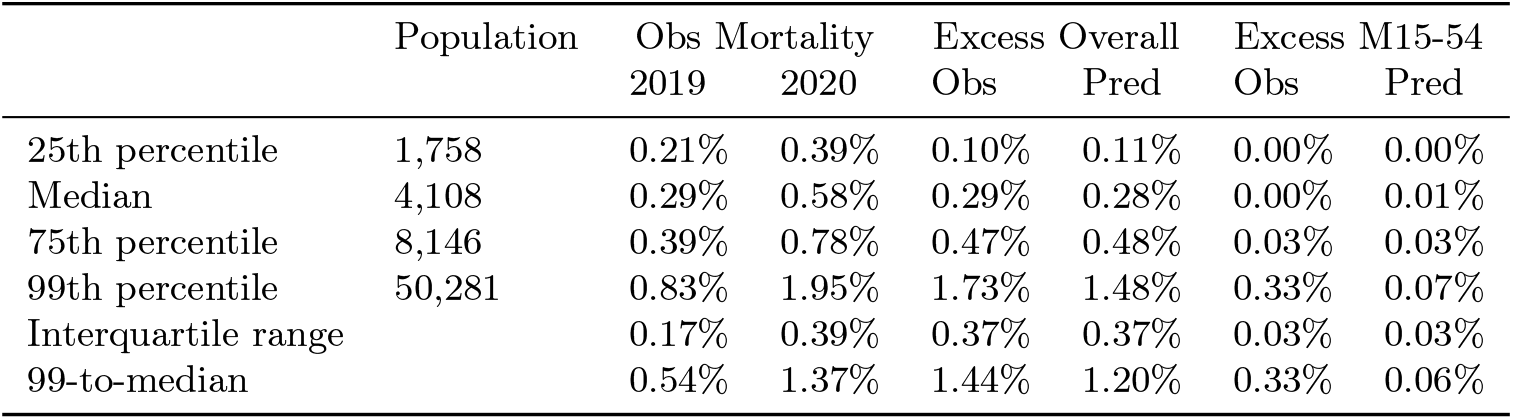
Summary Statistics for 612 Towns in Northern Italy

There is substantial variation in 2020 mortality across towns, and substantially more than in 2019. The interquartile range is 0.39 percentage points, versus 0.19 percentage points in 2019. This variation in observed mortality, which carries over to the excess mortality, is presumably due to the variation across towns in overall infection rates. This variation, and particularly the upper tail of the distribution of excess mortality across towns, is what we can exploit to calculate lower bounds for IFR.

Using the raw mortality for estimating IFR bounds by specific age and sex groupingsx presents problems, however. Some towns have low population and for groups with low underlying mortality rates, purely random variation will sometimes produce a small number of excess deaths. In towns with small populations such excess deaths translate into unusually high mortality *rates*, and these high rates will bias upwards the upper tail of the across-town distribution. Table 4 shows the problem. For the median town Males 15-54 have low overall mortality and zero excess mortality. In contrast, the upper tail (the 99th percentile) shows a high observed excess mortality of 0.33 percent. This is not a reliable estimate. The Poisson mixed effects model provides more reliable estimates.

Figures 1 and 2 shows the location of the towns. Figure 1 is a simple quantile map, showing towns by their predicted 2020 excess mortality. Figure 2 clusters towns by the excess mortality and contiguity (Euclidean distance). The maps indicate that mortality was high around Bergamo and particularly in the valleys north-east of Bergamo. Milan and surroundings appear to have been more lightly infected.

**Figure 1:**
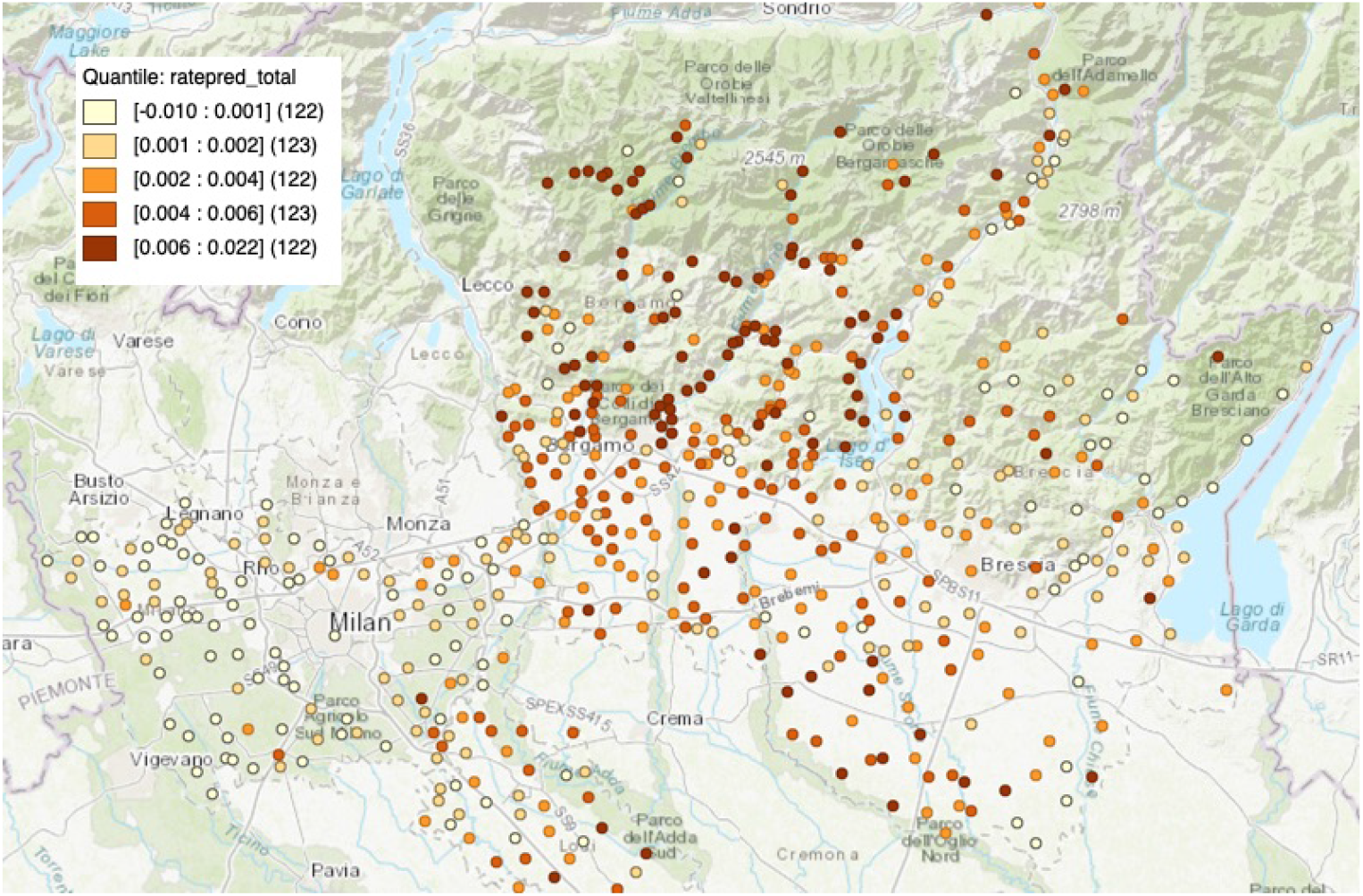
Quantile Map for Predicted Excess Mortality by Town in the Italian provinces of Milano, Bergamo, Brescia, and Lodi.

**Figure 2:**
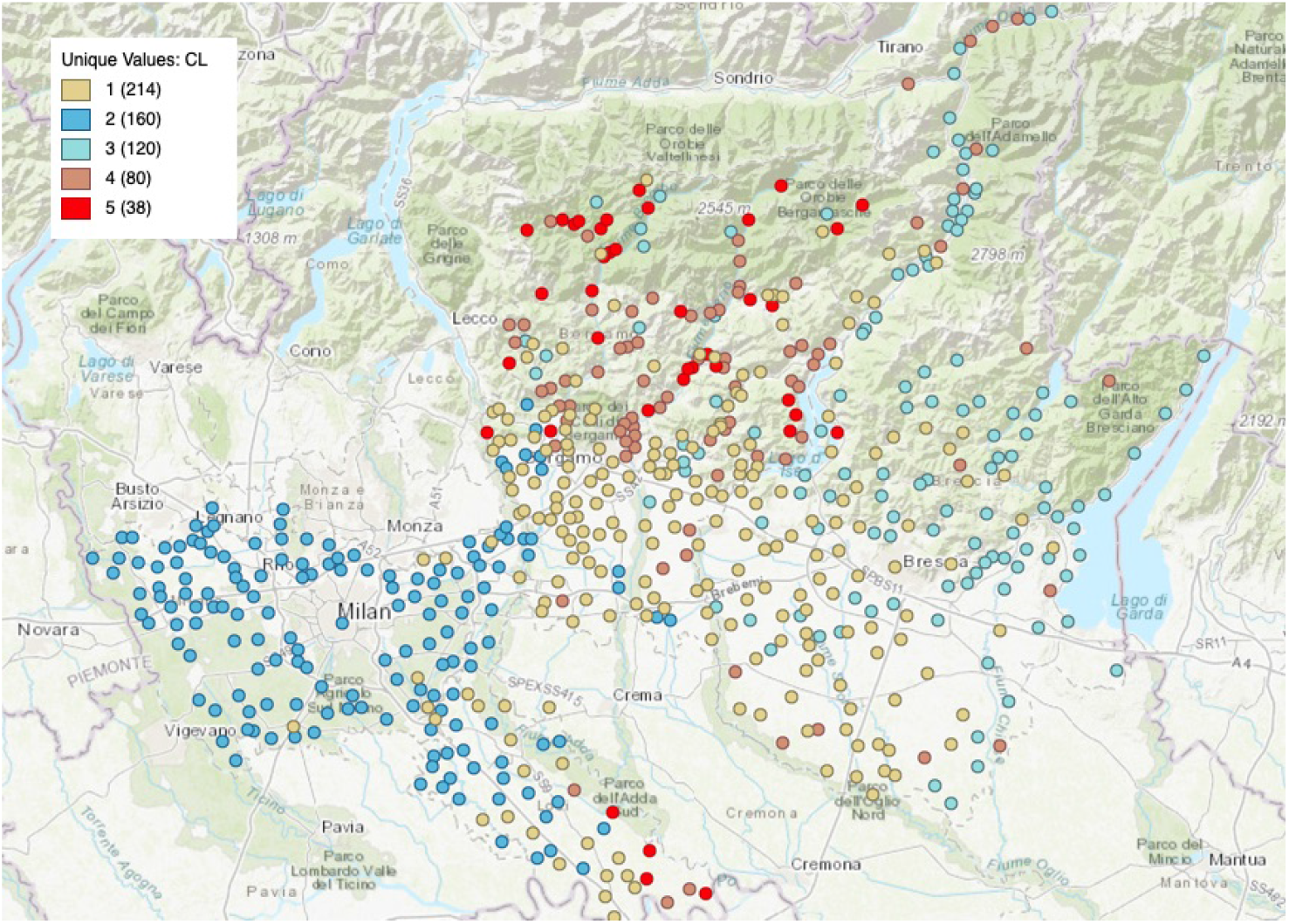
Clustered KMeans Map for Predicted Excess Mortality by Town in the Italian provinces of Milano, Bergamo, Brescia, and Lodi.

### 2.1 Data from ISTAT on Deaths in Italian Towns

ISTAT (Istituto Nazionale de Statistica, the Italian statistical agency) publish data on deaths by regions due to the coronavirus under https://www.istat.it/it/archivio/240401 – a cover page titled *Decessi e Cause di Morte: Cosa Produce L’ISTAT* (Deaths and Causes of Death: What ISTAT Produces). Under *Dataset analitico con i decessi giornalieri in ogni singolo comune di residenza* (Analytical dataset with daily deaths in each single municipality of residence (https://www.istat.it/it/files//2020/03/Dataset-decessi-comunali-giornalieri-e-tracciato-record.zip)) they report mortality “by gender and five-year age classes (for the first 4 months of the years from 2015 to 2019 for all 7904 municipalities in Italy, for the first three months of 2020 for 6,866 Municipalities and for the period from 1 January to 15 April 2020 for the 4,433 Municipalities verified in ANPR).” I aggregate this to

- Deaths from 1 January to 15 April for the years 2017, 2018, 2019, 2020^8^
- Deaths by five age groups: 0-14 years, 15-54, 55-64, 65-74, 75+
- Deaths for Male, Female, Total
- Select the provinces of Milano, Bergamo, Brescia, and Lodi which were particularly badly hit by the coronavirus, giving 612 towns

At http://demo.istat.it/pop2019/index3.html ISTAT reports population by town.^9^ In the region of Lombardy the four provinces of Milano, Bergamo, Brescia, and Lodi were particularly badly hit by the coronavirus. There are 612 towns with mortality data reported.

## 3 Poisson Count Mixed Effects Model

I use a mixed fixed and random effects Poisson count model (a form of general linear mixed model – see Raudenbush and Bryk [2002] Chapter 10 or Stroup [2013]). Data are grouped by:

- *i* – 612 towns
- *j* – ten demographic groups (Male 0-14, Male 15-54, Male 55-64, Male 65-74, Male 75+, Female 0-14, …)
- *t* – year (2017, 2018, 2019, 2020)

We have observations on:

Predicted excess mortality rate (from fixed effect estimation) clustered with GeoDa using KMeans clustering with 5 clusters, geometric centroids weighted at 0.33. The cluster center excess mortality (after standardizing) is: 1: 0.0038; 2: 0.0008, 3: 0.0008; 4: 0.0070; 5: 0.0126

- *Y*_*ijt*_ – deaths for each town, demographic group, and year for January 1 through April 15th for each of the years 2017-2020
- *POP*_*i jt*_ – population for each town, demographic group, for the years 2017-2019 (the 2019 population is used for 2020)
- *C*_*ijt*_ – COVID treatment (indicator variable for pre-2020 vs 2020)

This is a mixed effects model, with grouping by town (*i*) and demographic group (*j*), with the data being “treatment” of COVID versus pre-2020:

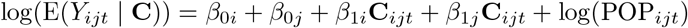

where the intercepts and slopes are specified as

*β* _0*i*_ = *α*_00_ + *α* _0*i*_ Town pre-2020 effect (intercept)

*β* _1*i*_ = *α*_10_ + *α* _1*i*_ Town 2020 effect (slope)

*β* _0*j*_ = *γ*_00_ + *γ* _0*j*_ Demographic pre-2020 effect (intercept)

*β* _1*j*_ = *γ*_10_ + *γ* _1*j*_ Demographic 2020 effect (slope)

I am going to treat all the *demographic* factors as fixed and (potentially) allow the *town* effects to be random. This seems reasonable because there are a large number of towns, each with a relatively small number of observations (eight demographic groups pre- and post-2020) but a small number of demographic groups each with a large number of observations (307 towns for each of pre- and post-2020). I can of course also examine this by treating all factors as fixed.) In this case the combined model is

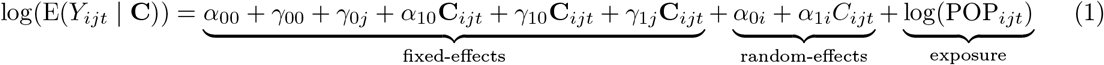

### Error Structure

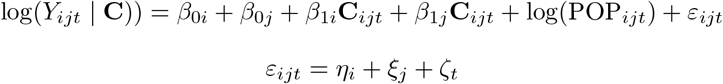

### Count, IFR, Infection Rate

By definition the number of deaths is:

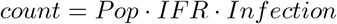

For now I am going to think about *mortality* rather than *IFR* and *Infection* separately:

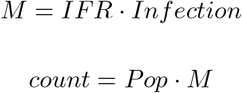

In Equation 1 the terms *α*_00_ + *γ*_00_ + *γ*_0*j*_ represent the pre-2020 age-specific mortality for the median town. The terms *α*_10_ + *γ*_10_ + *γ* _1*j*_ represent the age-specific excess mortality for the median town. The terms *α* _0*i*_ represent the random variation in pre-2020 town-specific mortality, and *α* _1*i*_ the 2020 town-specific excess mortality.

### 3.1 Fixed Effect versus Random Effect Estimators

The demographic effects are treated as fixed effects to be estimated:

- *α*_00_ + *γ*_00_ + *γ* _0*j*_ – 8 parameters (demographic groups) pre-COVID (pre-2020)
- *α*_10_ + *γ*_10_ + *γ* _1*j*_ – 8 parameters (demographic groups) for COVID mortality (2020)

The town effects (*α* _0*i*_ and *α* _1*i*_) can be treated as either fixed or random effects. There are 612 towns in the most-affected regions (Milano, Bergamo, Brescia, Lodi). The gives either 1,116x fixed effects (612 for for pre-2020 and 612 for 2020) or two random effects (one for pre-2020 and one for 2020).

The pre-2020 effects measures inherent base-case mortality differences across towns. (These are not due to different age and sex distributions since population and mortality are measured by the eight demographic categories listed above.) These pre-2020 effects are estimated using mortality from 2017, 2018, 2019. The 2020 effects provide for differences across towns in excess of their pre-2020 mortality. These mortality differences presumably result from different infection rates across towns, and possibly other factors such as the care and treatment provided locally.

Fixed effects estimators are appropriate for estimating specific town mortality, controlling for both pre-COVID mortality levels and noise in the 2020 mortality. Towns with low population will have low mortality counts and substantial variability in the counts. Across a large sample of towns some towns will produce large rates simply due to random variation. Randomly high counts for small towns will translate into high rates, potentially biasing any observations using raw rates.

Predicted mortality for town-specific rates using the fixed effects estimates will be more reliable than raw observations, particularly by demographic group. I am pooling age-specific counts across all towns to estimate age-specific rates, and pooling town-specific counts across age groups to estimate each town fixed effect. Standard errors for estimated fixed effects provide information on the confidence we should assign to town-specific estimates.

Random effects will be more appropriate for estimating the overall effect of COVID-19. The estimated pre-2020 age-specific mortality effects (*α* _0*i*_) provide estimates for the “average” town pre-COVID. The 2020 effects (*α* _1*i*_) estimate *excess* mortality for the “average” 2020 town - average in pre-2020 mortality and in 2020 infection rate.

## 4 Details on Estimation

Table 5 shows the coefficients for estimating Equation 1 as a mixed effect Poisson count model, with the town effects treated as random and the demographic effects as fixed. Using these estimates I can calculate predicted counts and thus predicted rates for the eight demographic groups both pre-COVID and for 2020. If we set the random effects at zero we will estimate excess mortality for the middle of the random effects distribution – i.e. the median town in terms of both pre-COVID and 2020 mortality. These results are shown in Table 8 below.^10^

**Table 5:**
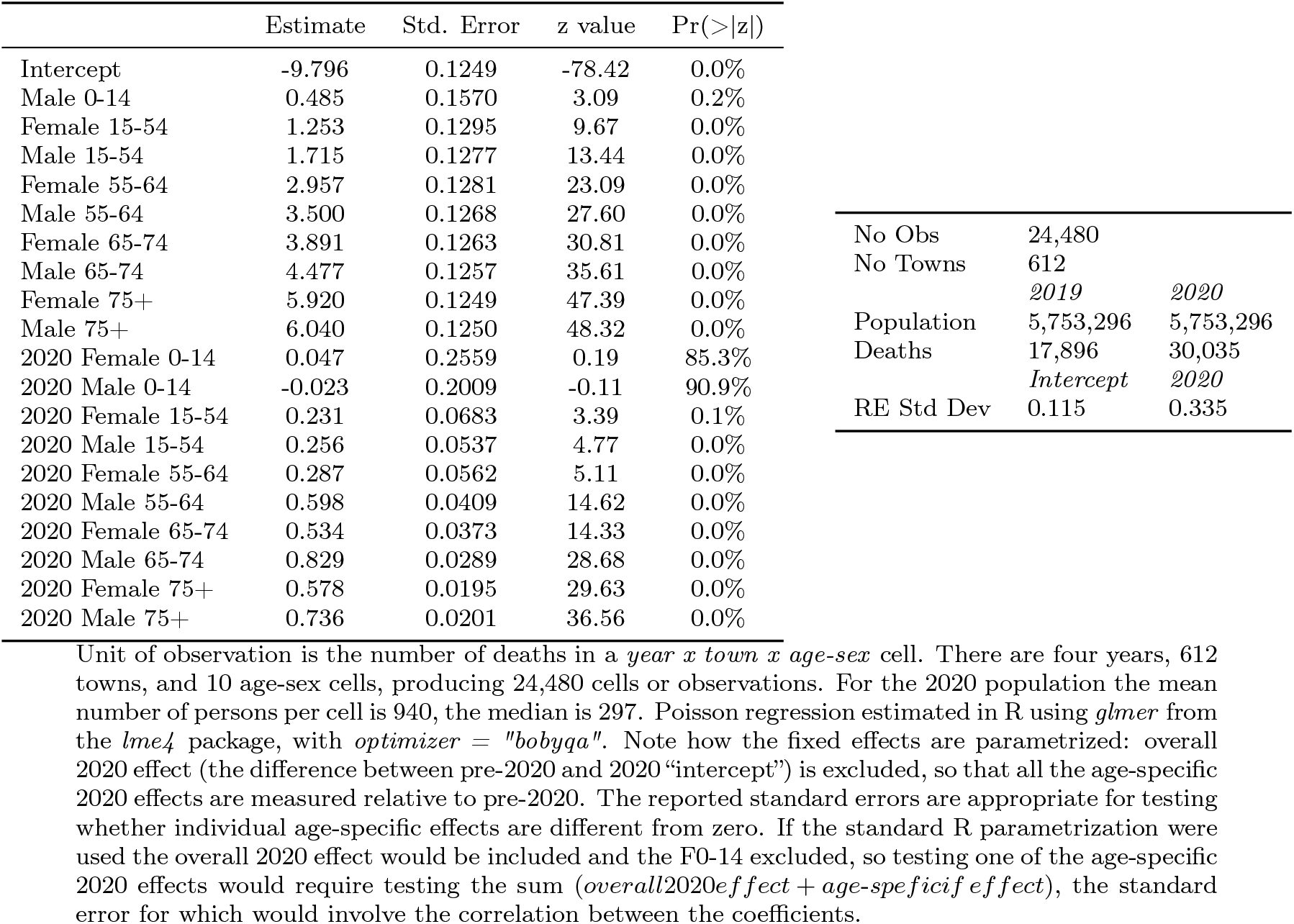
Estimated Random Effects Estimators (Town RE and Demographic FE)

We want the predicted excess mortality for the upper tail of the distribution – for towns that had high excess mortality and were thus, presumably, heavily infected. The estimated distribution of the 2020 random effect (*α* _1*i*_) measures this right tail (more infected towns) and will provide a lower bound for age-specific IFR. We use the 99th percentile of the *α* _1*i*_ distribution. If the estimated 2020 demographic fixed effect for a group *j* is 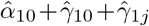 and the variance of the *α* _1*i*_ distribution is 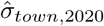,2020 then the 99th percentile (2.5*σ*) of the 2020 mortality will be larger than the pre-2020 mortality by the factor 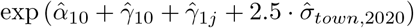. The excess mortality for group *j* will be

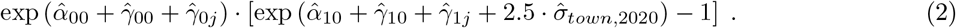

The coefficients in Table 5 show the age and sex profile of mortality in ratio terms (while the excess mortality in Table 1 show it as differences in rates – Equation 2). Mortality for demographic group *j* for the 99th-percentile town increased by the factor 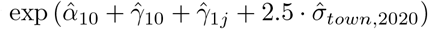. These increases, as a ratio to the pre-2020 mortality, are shown in Table 6; they are large and larger for older men.

**Table 6:**
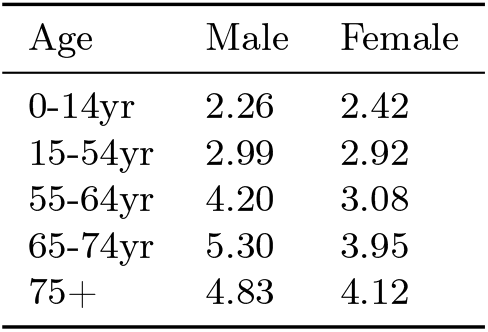
Fractional Increase in Mortality as a Ratio to pre-2020 Mortality

| Age | Male | Female |
| --- | --- | --- |
| 0-14yr | 2.26 | 2.42 |
| 15-54yr | 2.99 | 2.92 |
| 55-64yr | 4.20 | 3.08 |
| 65-74yr | 5.30 | 3.95 |
| 75+ | 4.83 | 4.12 |

Male mortality is higher than female pre-COVID at every age, with the ratio given by exp (*γ*_0 *F j*_ − *γ*_0 *Mj*_). The ratio exp (*γ*_1 *F j*_ − *γ*_1 *Mj*_) measures how much the ratio increases in 2020.

Table 7 shows summary statistics and test for the random and fixed effects models. I estimate three nested models for each the random and fixed effect specification. The first includes no estimates for 2020 effects – setting *α*_10_ + *γ*_10_ + *γ*_1 *j*_ and *α*_1 *i*_ to zero. The second includes town-specific effects, estimating *α*_1 *i*_. For both random and fixed effects models a likelihood ratio test shows that town-specific 2020 effects are important. The third model includes 2020 age-specific excess mortality effects – *α*_10_ + *γ*_10_ + *γ*_1 *j*_. Again, a likelihood ratio test shows that these effects are highly significant.

**Table 7:**
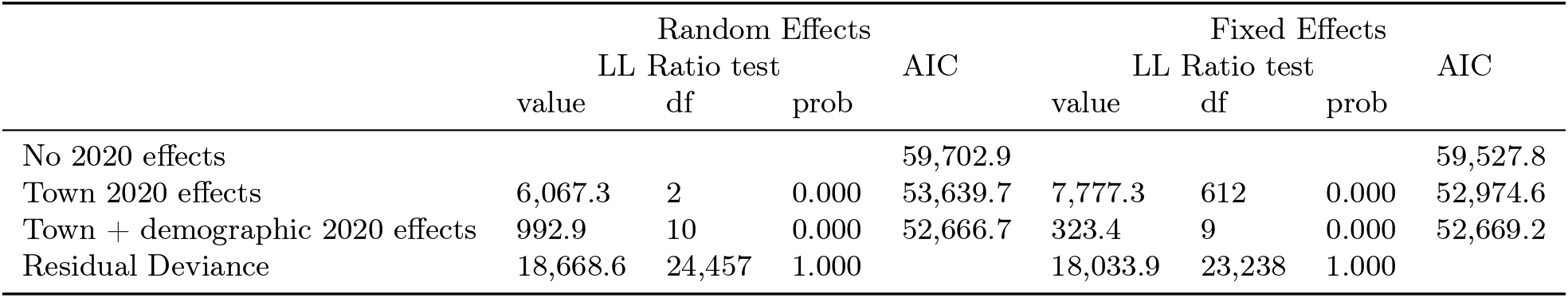
Summary Statistics for Random and Fixed Effects Estimation

The residual deviance in Table 7 provides a test for whether the Poisson regression adequately accounts for the observed variation – whether there is “overdispersion” relative to the Poisson assumption. The residual deviance is asymptotically *χ*^2^-distributed: a large value implies that the model does not account for the data very well. For both the random and the fixed effects model, the residual deviance is small relative to a *χ*^2^ random variable – the probability of observing a value as large or larger than reported is 1.0.

For reference, Table 8 shows the estimated excess mortality for the median town, towns with presumably modest overall infection rate.

**Table 8:** Estimated Excess Mortality, Middle of the Town Infection Distribution

| Age | Male | 95% CI | Female | 95% CI | Total | 95% CI |
| --- | --- | --- | --- | --- | --- | --- |
| 0-14yr | 0.00% | 0-0 | 0.00% | 0-0 | 0.00% | 0-0 |
| 15-54yr | 0.01% | 0.01-0.01 | 0.01% | 0-0.01 | 0.01% | 0-0.01 |
| 55-64yr | 0.15% | 0.13-0.17 | 0.04% | 0.02-0.05 | 0.09% | 0.08-0.1 |
| 65-74yr | 0.63% | 0.58-0.69 | 0.19% | 0.17-0.22 | 0.40% | 0.37-0.43 |
| 75+ | 2.54% | 2.4-2.71 | 1.62% | 1.51-1.72 | 1.97% | 1.87-2.09 |
| TOTAL | 0.32% | 0.3-0.34 | 0.26% | 0.24-0.28 | 0.29% | 0.28-0.31 |

Excess mortality rates calculated by eight age-sex categories then weighted to totals using age-specific population in the regions of Milano, Bergamo, Brescia, and Lodi. Non-parametric bootstrap confidence intervals.

**Table 9:** Percent of Population by Age and Sex, All Town 2020

| Age | Male | Female | Total |
| --- | --- | --- | --- |
| 0-14yr | 7.1% | 6.7% | 13.8% |
| 15-54yr | 25.9% | 25.0% | 50.9% |
| 55-64yr | 6.4% | 6.7% | 13.1% |
| 65-74yr | 5.0% | 5.6% | 10.6% |
| 75+ | 4.4% | 7.1% | 11.6% |
| TOTAL | 48.8% | 51.2% | 100.0% |

Calculating standard errors for the excess mortality is difficult. For a specific age group this would be the 99th percentile 2020 rate less the base-case pre-2020 rate shown in Equation 2. Standard errors for that expression is not easy because it is a non-linear expression of the coefficients. One might try doing so by the delta-method but that is complicated by two factors: first the standard error for 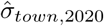 is not easy to obtain, and second we want to aggregate age- and sex-specific rates to overall rates using regional population weights, leading to weighted averages of terms like Equation 2. The most feasible alternative is bootstrapping. Non-parametric bootstrapping where towns are sampled from the original distribution and Equation 1 re-estimated for each sample. Tables 1 and 8 show the results for bootstrapping with 400 samples (sampling with replacement from the town distribution).

Figure 3 shows a histogram of estimated 2020 excess mortality across the 612 towns. This shows how towns varied in their 2020 excess mortality, presumably resulting from different coronavirus infection rates. The distribution is skewed with a longer right tail. Figure 4 shows the estimated 2020 fixed effects across towns. The fixed effects show a more symmetric distribution than do the predicted excess mortality, because the fixed effects are measured in log terms (see Equation 1). The fixed effects correspond to the random effects in Equation 1 and the more symmetric distribution of the estimated fixed effects provides some support for the assumption that the random effects are normally distributed.

**Figure 3:**
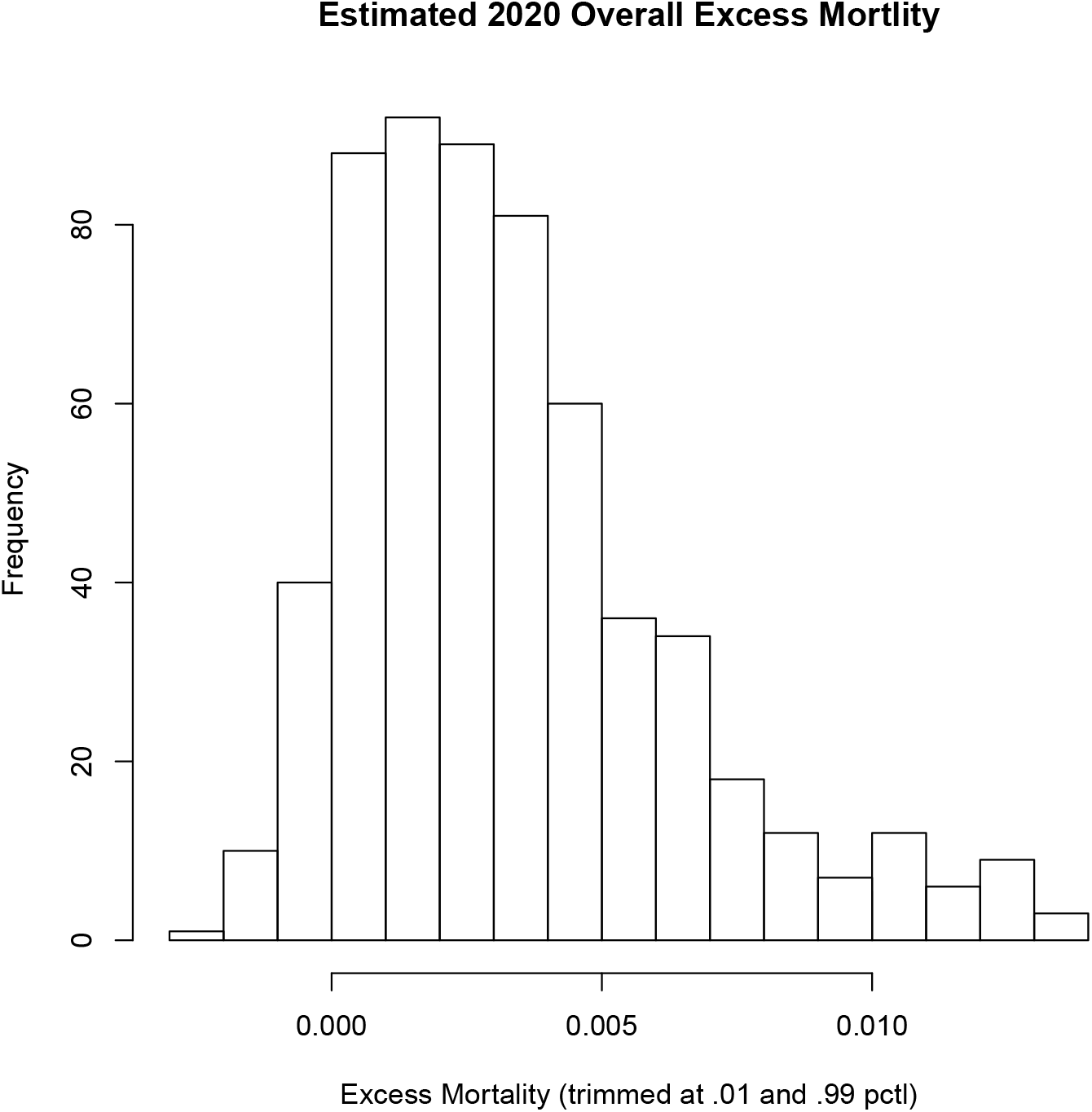
Histogram of Predicted Excess Mortality by Town.

**Figure 4:**
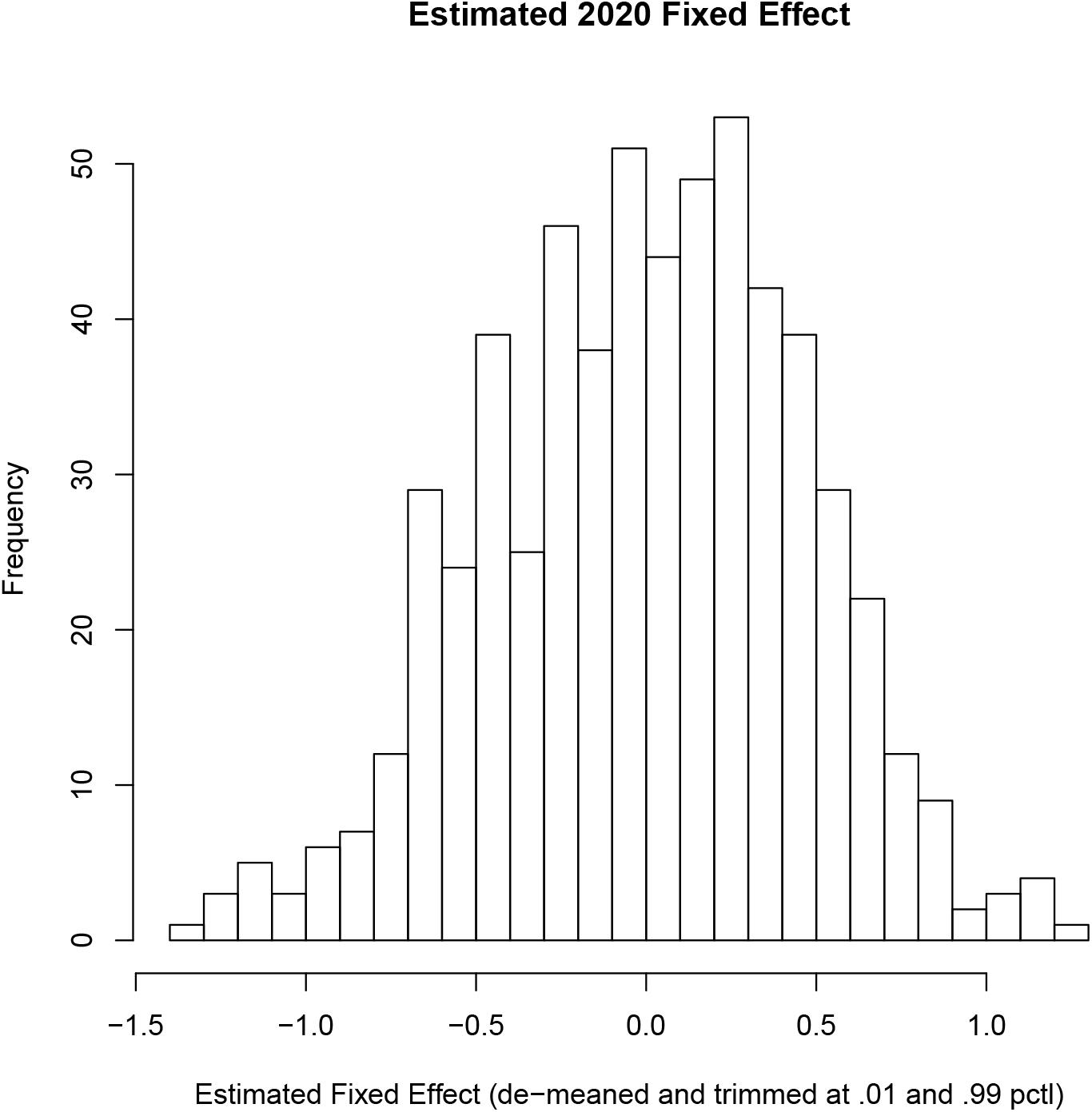
Histogram of Estimated Town Fixed Effect.

## Data Availability

All mortality and population data are publicly available from ISTAT. I have also published the code and data on GitHub

https://www.istat.it/it/archivio/240401

http://demo.istat.it/pop2019/index3.html

https://github.com/tscoleman/COVID_NorthernItalianTowns

## Acknowledgements

Agostino Russo proposed the original idea of using mortality data from ISTAT for the towns of Nembro and Alzano as lower bounds for the IFR. Xi Song corrected my notation. Errors are my own.

## A Version History

**10-jun-20** V1 – initial submission

**12-jun-20** V2 – in Table 2 correct “Total” resulting from error in population weights; add histogram of estimated fixed effects.

## Footnotes

1 The rates by age and sex are estimated from the underlying data, and then aggregated to the totals using population weights for the overall region (shown in Table 9).

2 The ratio of male-to-female

3 There is a fourth possible source for the observed increase in mortality: some unobserved confounding factor that increased mortality in northern Italian towns in spring 2020. Although logically possible this is unlikely to be a major factor in this case, particularly given two observations: the clear identification of a mechanism (virus and disease) causing increased mortality globally, and significant increases in mortality in northern Italian towns that occurred at the same time as identified COVID-19 infection and in tandem in all groups except the youngest.

4 The CDC scenario 3 has the highest implied IFR of 0.8 percent overall and 2.56 percent for 65+, with the overall somewhat lower than our lower bounds.

5 The age-specific mortality is predicted for each town, then weighted using that town’s population weights.

6 The predicted overall morality from the random effects estimation shown in Table 1 is lower than for these towns at the 99th percentile. This effect, shrinkage of the random effects distribution, is a characteristic of mixed models – see Clark [2019].

7 Observed excess mortality is calculated as the difference between the 2020 mortality and 2019 mortality on a town-by-town basis, so the median of the excess mortality may not equal the difference in the medians of 2020 and 2019. The Predicted excess mortality is calculated from the Fixed Effects estimator as 2020 predicted mortality versus the pre-2020 mortality estimated using 2017, 2018, 2019.

8 The ISTAT data extract includes deaths through 30 April for the years before 2020, so observations for days after 15 April are excluded.

9 Earlier years are accessed by replacing “2019” with “2018” etc.

10 Using the observed population for all provinces combined (weights for 2020 shown in Table 9, total population reported in Table 5) produces predicted counts for all provinces combined. The difference in counts provides an estimate of the excess mortality by demographic groups. Summing counts across male and female gives the population-weighted averages, and dividing by the population gives the population-weighted excess mortality

## References

CDC. Coronavirus Disease 2019 (COVID-19), February 2020. URL https://www.cdc.gov/coronavirus/2019-ncov/hcp/planning-scenarios.html. Library Catalog: www.cdc.gov.

Michael Clark. Mixed Models in R, February 2019. URL https://m-clark.github.io/mixed-models-with-R/.m-clark.github.io.

Giacomo Grasselli, Antonio Pesenti, and Maurizio Cecconi. Critical Care Utilization for the COVID-19 Outbreak in Lombardy, Italy: Early Experience and Forecast During an Emergency Response. JAMA, 323(16):1545–1546, April 2020. ISSN 0098-7484. doi: 10.1001/jama.2020.4031. URL https://jamanetwork.com/journals/jama/fullarticle/2763188. Publisher: American Medical Association.

Richard Grewelle and Giulio De Leo. Estimating the Global Infection Fatality Rate of COVID-19. medRxiv, page 2020.05.11.20098780, May 2020. doi: 10.1101/2020.05.11.20098780. URL https://www.medrxiv.org/content/10.1101/2020.05.11.20098780v1. Publisher: Cold Spring Harbor Laboratory Press.

John Ioannidis. The infection fatality rate of COVID-19 inferred from seroprevalence data. medRxiv, page 2020.05.13.20101253, May 2020. doi: 10.1101/2020.05.13.20101253. URL https://www.medrxiv.org/content/10.1101/2020.05.13.20101253v1. Publisher: Cold Spring Harbor Laboratory Press.

Davide Manca. Dynamics of ICU patients and deaths in Italy and Lombardy due to Covid-19 Analysis updated to 30-March, Day #38 evening. Technical report, European Society of Anesthesiology, March 2020. URL https://www.esahq.org/esa-news/dynamics-of-icu-patients-and-deaths-in-italy-and-lombardy-due-to-co Library Catalog: www.esahq.org Publisher: ESA HQ.

Stephen W. Raudenbush and Anthony S. Bryk. Hierarchical linear models: applications and data analysis methods. Sage Publications, Thousand Oaks, 2nd ed. edition, 2002. ISBN 978-0-7619-1904-9. URL http://pi.lib.uchicago.edu/1001/cat/bib/4649191.

Walter W. Stroup. Generalized linear mixed models: modern concepts, methods and applications. Chapman & Hall/CRC texts in statistical science series. CRC Press, Boca Raton, FL, 2013. ISBN 978-1-4398-1512-0.

